# Develop and Validate a Computable Phenotype for the Identification of Alzheimer’s Disease Patients Using Electronic Health Record Data

**DOI:** 10.1101/2024.02.06.24302389

**Authors:** Xing He, Ruoqi Wei, Yu Huang, Zhaoyi Chen, Tianchen Lyu, Sarah Bost, Jiayi Tong, Lu Li, Yujia Zhou, Jingchuan Guo, Huilin Tang, Fei Wang, Steven DeKosky, Hua Xu, Yong Chen, Rui Zhang, Jie Xu, Yi Guo, Yonghui Wu, Jiang Bian

## Abstract

**INTRODUCTION:** Alzheimer’s Disease (AD) are often misclassified in electronic health records (EHRs) when relying solely on diagnostic codes. This study aims to develop a more accurate, computable phenotype (CP) for identifying AD patients by using both structured and unstructured EHR data.

**METHODS:** We used EHRs from the University of Florida Health (UF Health) system and created rule-based CPs iteratively through manual chart reviews. The CPs were then validated using data from the University of Texas Health Science Center at Houston (UT Health) and the University of Minnesota (UMN).

**RESULTS:** Our best-performing CP is “*patient has at least 2 AD diagnoses and AD-related keywords*” with an F1-score of 0.817 at UF, and 0.961 and 0.623 at UT Health and UMN, respectively.

**DISCUSSION:** We developed and validated rule-based CPs for AD identification with good performance, crucial for studies that aim to use real-world data like EHRs.

## BACKGROUND

Alzheimer’s disease (AD) and AD-related dementias (AD/ADRD) represent complex neurodegenerative diseases affecting approximately 6.7 million Americans over 65 and over 40 million people worldwide.^1–3^ Significant efforts have been made to better understand AD/ADRD, seek effective treatments and prevention strategies, and address the needs of AD/ADRD patients. The United States (US) National Alzheimer’s Project Act (NAPA) has recommended a $2 billion annual budget and calls for an aggressive and coordinated national plan for accelerating AD/ADRD research and improving patient care.^4^

The widespread adoption of electronic health record (EHR) systems has made large-scale, longitudinal clinical data available for research. As an important real-world data (RWD) source,^5^ EHR have become increasingly important for generating real-world evidence^6^ in AD/ADRD research reflecting the patient population treated in real-world clinical settings. For example, Miller et al. examined the prevalence of AD/ADRD in the state of Florida and characterize the demographic characteristics of the AD/ADRD population using EHR data from the OneFlorida (now OneFlorida+) Clinical Research Consortium.^7^ Many studies have also developed AD/ADRD prediction models using diagnosis, medication history, and biomarker data from RWD like EHRs and administrative claims.^8–11^ However, to effectively utilize RWD in AD/ADRD research, a key prerequisite is to accurately identify the target populations, especially when AD/ADRD onset are used as outcome labels. Previously, AD/ADRD cohorts were often identified solely by diagnostic codes (e.g., International Classification of Diseases [ICD]), leading to significant misclassification errors. High variations in classification accuracies have been reported in validation studies for using diagnostic codes to define dementia including AD/ADRD.^12^ In a study that used two Swedish national RWD registers and six population-based studies, Feldman et al. found that relying solely on diagnostic codes yields a positive predictive value (PPV) of only 0.82. In two other studies using Danish nationwide hospital registers, diagnostic codes accurately identified AD in only 60-80% of cases, with a PPV ranging from 0.78 to 0.81.^13,14^ In a US based study, Taylor et al. found that the AD diagnostic codes in Medicare claims data only have a sensitivity of 0.64 and a specificity of 0.96 for identifying AD.^15^ Besides diagnostic codes, EHR data element like AD-related medications have also been used to identify AD patients. Tjandra et al. developed and validated an AD cohort discovery tool using a rule set that includes encounters, diagnosis codes, medications, and procedure codes (e.g., for psychological/cognitive testing), achieving moderate performance with an F1-score of 0.73, a PPV of 0.77, and a sensitivity of 0.70 in a Michigan Alzheimer’s Disease Research Center (ADRC) cohort.^16^

Identifying patients with a particular condition, e.g., AD, within the context of EHRs, is accomplished through a computable phenotype (CP) or simply phenotype (traditionally often called cohort identification or case finding algorithms), which is defined “*clinical conditions, characteristics, or sets of clinical features that can be determined solely from EHRs and ancillary data sources and does not require chart review or interpretation by a clinician.*”^17^ CPs have gained popularity for their high specificity and sensitivity in EHR-based cohort identification, showing success in various domains, including the identification of HIV prevalent cases, transgender and gender nonconforming individuals, resistant hypertension, among others.^18–20^ Traditionally, EHR-based CPs only considered structured information (e.g., diagnoses, medications, etc.), while EHR also contains rich unstructured clinical narratives (e.g., progression notes, discharge summaries).^21^ In fact, over 80% of patient information in EHR is documented in free-text clinical narratives,^21^ which contain more detailed patient information including important variables such as cognitive assessments that can facilitate the identification of AD patients. Prior studies in other disease domains that leverage both structured EHR data and unstructured narratives can also significantly improve the performance of CPs.^22,23^

In this study, we developed and validated a CP to accurately identify individuals with AD using both structured and unstructured data from University of Florida Health (UFHealth). Additionally, we assessed the prevalence of AD patients within UFHealth from the identified cohort and reported patient characteristics.

## METHODS

### Data sources

We retrieved individual patient-level data from the UFHealth Integrated Data Repository (IDR) after obtaining approval from the UF Institutional Review Board (IRB). The UFHealth IDR serves as an enterprise data warehouse consisting of data across UFHealth’s clinical and administrative information systems (e.g., Epic electronic health record [EHR] system), covering a population of over 1 million patients^24^. We then used data from the University of Texas Health Science Center at Houston (UTHealth) Physicians Clinical Data Warehouse (CDW) and the University of Minnesota (UMN) Academic Health Center Information Exchange (AHC-IE) clinical data repository (CDR) for external validation of the CPs developed using UFHealth data. The UTHealth Physicians CDW encompasses all UTHealth Physicians outpatient EHR data, serving approximately 1.8 million patients. And the UMN AHC-IE CDR comprises data from over 4.5 million patients who received care at 8 hospitals and more than 40 clinics.

### Overall study design

We developed the CP for identifying AD patients by using both structured and unstructured EHR data. As shown in ***Figure 1***, we adopted a two-step process to develop the AD CP: (1) we first identified a potential AD cohort via searching EHRs for patients with at least one AD-related diagnosis codes, i.e., International Classification of Diseases-Nineth/Tenth Revision-Clinical Modification (ICD-9/10-CM) codes as shown in **Table 1**. For all the patients within the cohort, we collected their EHR data, including structured data (e.g., demographic, diagnoses, procedures, medications, laboratory results, procedures, etc.) and unstructured clinical notes (e.g., progress notes, discharge summaries, pathology reports, etc.); and (2) we then iteratively derived the CP rules through manual chart reviews on selected samples from the potential AD cohort.

**Figure 1.**
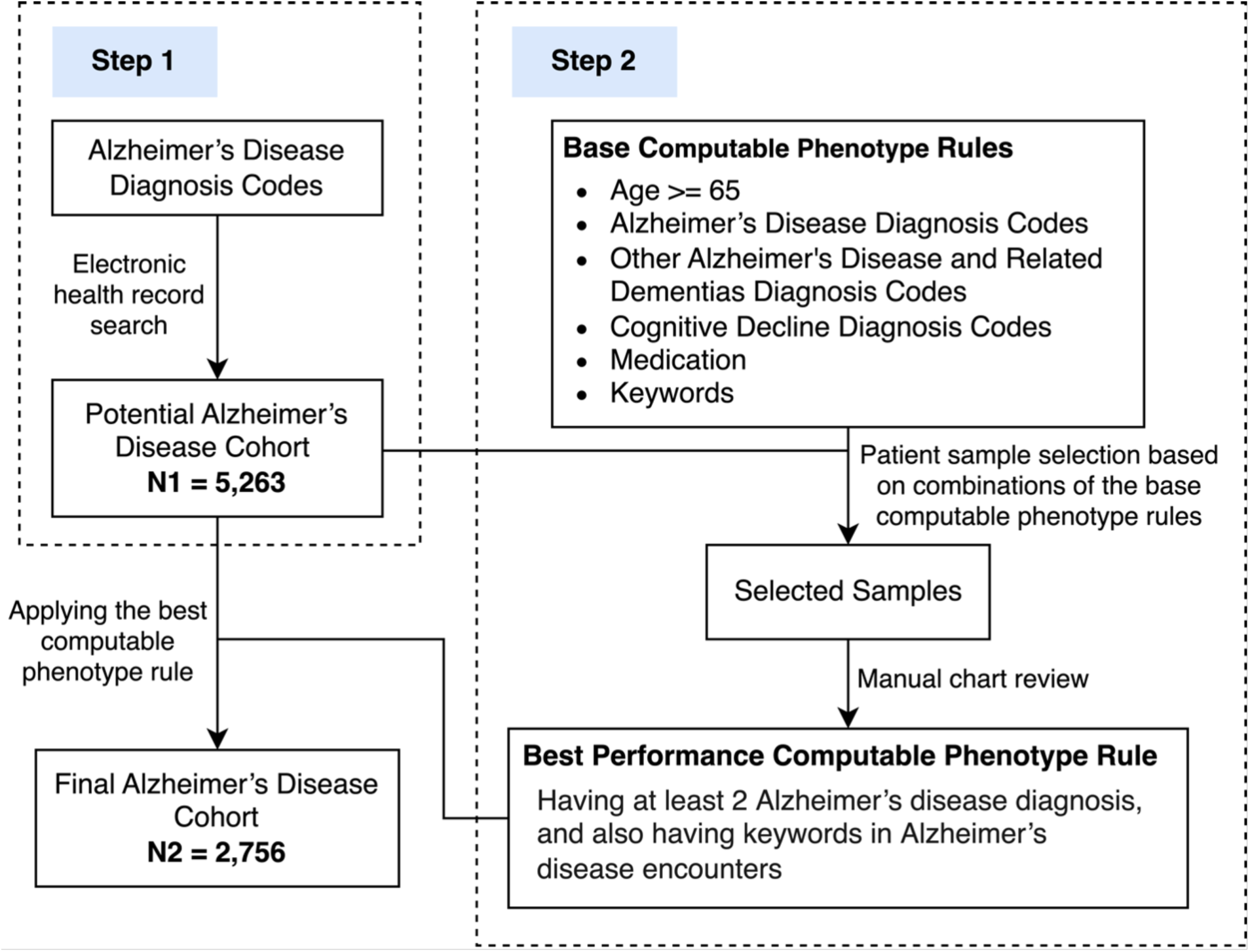
Flow chart of the Alzheimer’s’ disease computable phenotype development process.

**Table 1.** Value sets, codes, and keywords used in the 7 base rules.

|  | **Value Sets/Codes/Keywords** |
| --- | --- |
| **AD Diagnosis Codes** |  |
| ICD-9-CM | 331.0 – “Alzheimer's disease” |
| ICD-10-CM | G30 – “Alzheimer's disease”<br>G30.0 – “Alzheimer's disease with early onset”<br>G30.1 – “Alzheimer's disease with late onset”<br>G30.8 – “Other Alzheimer's disease”<br>G30.9 – “Alzheimer's disease, unspecified” |
| **Other ADRD Diagnosis Codes (Selected Examples)** |  |
| ICD-9-CM | *e.g., 290.0 – “Senile dementia, uncomplicated”, 331.11 – “Pick's disease”, 437.2 – “Hypertensive encephalopathy”, etc.* |
| ICD-10-CM | *e.g., F01.50 – “Vascular dementia without behavioral disturbance”, G91.0 – “Communicating hydrocephalus”, G94 – “Other disorders of brain in diseases classified elsewhere”, etc.* |
| **Cognitive Decline Diagnosis Codes (Selected Examples)** |  |
| ICD-9-CM | *e.g., 78093 – “Memory loss”, 79952 – “Cognitive communication deficit”, 331.83 – “Mild cognitive impairment, so stated”, etc.* |
| ICD-10-CM | *e.g., G31.84 – “Mild cognitive impairment, so stated”, G31.89 – “Other specified degenerative disease of nervous system”, R41.81 – “Age-related cognitive decline”, etc.* |
| **Medication (Selected Examples)** |  |
| Names/RxNorm | *e.g., “Aricept”, “Namzaric Oral Product”, “24 HR galantamine hydrobromide 16 MG Extended Release Oral Capsule”, “Namenda”, etc.* |
| **Keywords (Selected Examples)** |  |
|  | *e.g., “dementia”, “Alzheimer”, “memory loss”, “cognitive deficits”, “cognitive impairment”, “cognitive disorders”, “cognitive decline”, “amnesia”, etc.* |

### Derive CP rules based on manual chart reviews on selected samples from the potential AD cohort

Based on insights from previous studies on case finding algorithms for AD^16^ and dementia^11,12,25,26^, as well as consultations with clinicians who specialize in AD patient care, we proposed 7 initial base rules: (1) age equal to or larger than 65, (2) having at least 2 of the AD diagnosis codes, (3) having at least 5 of the AD diagnosis codes, (4) having at least 1 of the other ADRD diagnoses, (5) having at least 1 of the cognitive decline diagnosis, (6) having at least 1 of the relevant medication use, and (7) having at least 1 of the relevant keywords in notes from AD encounter. The value sets, codes, and keywords used in the 7 base rules are listed in ***Table 1***.

We generated a total of 69 distinct combinations by combining these 7 base rules, as shown in ***Table 2***. Subsequently, we randomly sampled 10% of the patients who met the rule combination for each of these 69 combinations. If the number of patients for the 10% sample is larger than 20, we employed random selection to pick 20 patients for this combination. Conversely, if the total patient count for a combination was less than 3, we manually reviewed all patients within that combination. In total, we selected 363 patients and split them into a training set and a testing set in an 8:2 ratio (i.e., 282 for training samples and 81 for testing samples). Only the training set was used to develop the CP rules.

**Table 2.** Summary of Top 8 combinations of base rules and the number of patients who identified by each rule combination and the number of actual patients confirmed by manual chart reviews.

| Age | AD diagnosis Codes |  | Other AD/DRD diagnosis Codes | Cognitive decline diagnosis Codes | Medication | Keyword | Total # of Patients<sup>a</sup> | # of Patients Selected | # of AD Patients<sup>b</sup> |
| --- | --- | --- | --- | --- | --- | --- | --- | --- | --- |
| ≥65 | ≥2 | ≥5 | ≥1 | ≥1 | ≥1 | ≥1 |  |  |  |
| + | - | - | + | - | + | + | 773 | 20 | 2 |
| + | - | - | + | - | - | + | 616 | 20 | 1 |
| + | + | + | + | + | + | + | 565 | 20 | 19 |
| + | + | - | + | - | + | + | 554 | 20 | 11 |
| + | + | - | + | + | + | + | 552 | 20 | 17 |
| + | - | - | + | + | + | + | 430 | 20 | 0 |
| + | + | + | + | - | + | + | 283 | 20 | 17 |
| + | + | - | + | - | - | + | 235 | 20 | 12 |
| Note that ‘+’ indicates that the patient must meet this rule.<br><sup>a</sup>Overall, there were 69 different rule combinations that identified patient. Only the top 8 rule combinations are displayed here. All the 69 rule combinations are reported in **Supplement Table 1**.<br><sup>b</sup>Number of AD patients identified by chart review from the selected patient sample. |  |  |  |  |  |  |  |  |  |

We first developed an annotation guideline for the manual chart reviews to ensure consistent criteria were applied across all reviewers. Three annotators (TL, PY, and SB) conducted the chart review iteratively in rounds. In each round, they individually reviewed the same 10 samples from the training set following the annotation guideline. After each round, if any disagreements arose among the three reviewers, the entire study team engaged in discussions to resolve these conflicts and reach a consensus and the annotation guideline was iteratively revised accordingly to these discussions. After 5 rounds of chart reviews (i.e., after assessing 50 samples), the inner-rater agreements between any two of the three annotators achieved a Cohen’s kappa of 1, indicating perfect agreement. Subsequently, the three annotators began annotating the remaining training samples independently, with the explicit instruction to be cautious when a case was deemed ambiguous. In cases where inconsistent annotations were encountered, a fourth reviewer (JB) helped in resolving the discrepancies. We assessed the performance of each rule combination on both the training and testing sets using multiple metrics, including specificity, sensitivity, positive predictive value (PPV), negative predictive value (NPV), and F1-score. The rule with the highest F1-score was selected as the final CP.

### External validation

To further validate the CP and evaluate its generalizability, we applied the CP rules on EHR data from UTHealth and UMN. We conducted manual chart reviews of a random sample of 50 patients from each site using the same annotation guideline used at UFHealth.

## RESULTS

### Development of the computable phenotype for the identification of AD patients

Using the AD diagnosis codes (i.e., ICD codes in ***Table 1***), we identified a potential AD cohort consisting of 5,263 patients from the UFHealth IDR. Among this cohort, our final CP identified 2,756 patients as AD patients, as shown in ***Figure 1***. A final set of CP algorithms was selected based on the best F1-score of the various base CP rule combinations listed in ***Table 3***, using manual chart review results as the gold standard. The best performing CP with structured data only was “*the patient has at least 2 AD diagnoses*”, having an F1-score of 0.787 on the training set. When considering both structured and unstructured data, the best performing CP was “*the patient has at least 2 AD diagnoses and has keywords in AD encounters*”, with an F1-score of 0.829 outperforming the one considering structured data only. The performance of these CP algorithms was further assessed using the independent testing set (i.e., the test sample with 81 patients). When applying the final CP algorithms to the testing set, the CP with structured data received an F1-score of 0.789, while the CP using both structured and unstructured data achieved a better F1-score of 0.817. Both CP rules exhibited significant improvements in F1-scores compared to the baseline CP rule. ***Table 3*** shows the different performance metrics (i.e., sensitivity, PPV, and F1-score) of these CP algorithms under different settings.

**Table 3.** The performance of the baseline CP rule and the best performing CP rules in terms of F1-score on the training data and the testing data.

|  | Rule<sup>a</sup> | Sensitivity | PPV | F1-score |
| --- | --- | --- | --- | --- |
| **Training Data** |  |  |  |  |
| Baseline | with at least 1 AD diagnoses | 1.000 | 0.415 | 0.586 |
| Structured Data Only (best F1-score) | with at least 2 AD diagnoses | 0.949 | 0.673 | 0.787 |
| Structured and Unstructured Data (best F1-score) | with at least 2 AD diagnoses, and with keywords in AD encounters | 0.932 | 0.747 | 0.829 |
| **Testing Data** |  |  |  |  |
| Baseline | with at least 1 AD diagnoses | 1.000 | 0.370 | 0.541 |
| Structured Data Only (best F1-score) | with at least 2 AD diagnoses | 1.000 | 0.652 | 0.789 |
| Structured and Unstructured Data (best F1-score) | with at least 2 AD diagnoses, and with keywords in AD encounters | 0.967 | 0.707 | 0.817 |

### External validation

There are 6,821 patients and 10,387 patients with at least one AD diagnosis code were identified at UTHealth and UMN sites, respectively. For the validation process, a random sample of 50 patients from each site were selected for manual chart reviews. ***Table 4*** shows the performance of the final CP rules on the two validation sites. Our CPs showed different performance across the two sites. For the structured data only CP, the F1-scores were 0.871 and 0.667, respectively, for the two validation sites. On the other hand, the CP using both structured data and unstructured data had F1-scores of 0.961 and 0.623 for the respective sites.

**Table 4.** The performance of final CP rules on external validation sites.

|  | Rule | Sensitivity | PPV | F1-score |
| --- | --- | --- | --- | --- |
| **UTHealth validation data** |  |  |  |  |
| Structured Data Only | with at least 2 AD diagnoses | 0.974 | 0.787 | 0.871 |
| Structured and Unstructured Data | with at least 2 AD diagnoses, and with keywords in AD encounters | 0.974 | 0.949 | 0.961 |
| **UMN validation data** |  |  |  |  |
| Structured Data Only | with at least 2 AD diagnoses | 0.550 | 0.846 | 0.667 |
| Structured and Unstructured Data | with at least 2 AD diagnoses, and with keywords in AD encounters | 0.475 | 0.905 | 0.623 |

### Definitive AD cohort characteristics

We applied the best performing CP using both structured and unstructured data (i.e., “*the patient has at least 2 AD diagnoses and has keywords in AD encounters*”). ***Table 5*** describes the patient characteristics for both the definitive AD cohort and the potential AD cohort across the three sites. Compared with the potential AD cohort, the definitive AD cohort is slightly younger with more females and non-Hispanic Blacks. In addition to demographics, we also examined the prevalence of several chronic conditions in these cohorts. The definitive AD cohort shows a higher prevalence of patients with depression, hypertension, and cancer compared to the potential AD cohort.

**Table 5.** Characteristics of the definitive AD cohort and the potential AD cohort within UFHealth IDR, UTHealth Physicians CDW, and UMN AHC-IE CDR.

|  | UFHealth IDR |  | UTHealth Physicians CDW |  | UMN AHC-IE CDR |  |
| --- | --- | --- | --- | --- | --- | --- |
|  | Definitive AD cohort<sup>a</sup> | Potential AD cohort<sup>b</sup> | Definitive AD cohort | Potential AD cohort | Definitive AD cohort | Potential AD cohort |
|  | N = 2,756 | N = 5,263 | N = 1,823 | N = 6,821 | N = 5,311 | N = 10,387 |
| **Age** |  |  |  |  |  |  |
| < 55 | 53 (1.9%) | 111 (2.1%) | 124 (6.80%) | 433 (6.35%) | 52 (1.0%) | 105 (1.0%) |
| 55-64 | 194 (7.0%) | 364 (6.9%) | 327 (17.94%) | 981 (14.38%) | 170 (3.2%) | 354 (3.4%) |
| 65-74 | 665 (24.1%) | 1,165 (22.1%) | 607 (33.30%) | 2,011 (29.48%) | 778 (14.6%) | 1,347 (13.0%) |
| 75-84 | 1,178 (42.7%) | 2,183 (41.5%) | 620 (34.01%) | 2,472 (36.24%) | 2,136 (40.2%) | 3,939 (37.9%) |
| >= 85 | 666 (24.2%) | 1,440 (27.4%) | 144 (7.90%) | 911 (13.36%) | 2,175 (41.0%) | 4,642 (44.7) |
| Unknown | 0 (0%) | 0 (0%) | 1 (0.05%) | 13 (0.19%) | 0 (0%) | 0 (0%) |
| **Sex** |  |  |  |  |  |  |
| Male | 1,028 (37.3%) | 2,023 (38.4%) | 669 (36.70%) | 2,490 (36.50%) | 1,783 (33.6%) | 3,620 (34.9%) |
| Female | 1,728 (62.7%) | 3,240 (61.6%) | 1,152 (63.19%) | 4,317 (63.29%) | 3,528 (66.4%) | 6,766 (65.1%) |
| Unknown | 0 (0%) | 0 (0%) | 2 (0.11%) | 14 (0.21%) | 0 (0%) | 0 (0%) |
| **Race/Ethnicity** |  |  |  |  |  |  |
| Hispanics | 148 (5.4%) | 239 (4.5%) | 59 (3.24%) | 492 (7.21%) | 50 (0.9%) | 98 (0.9%) |
| NHW | 1,828 (66.3%) | 3,648 (69.3%) | 1,011 (55.46%) | 2,883 (42.27%) | 4,681 (88.1%) | 9,253 (89.1%) |
| NHB | 643 (23.3%) | 1,094 (20.8%) | 323 (17.72%) | 1,548 (22.69%) | 142 (2.7%) | 275 (2.6%) |
| Other | 90 (3.3%) | 136 (2.6%) | 291 (15.96%) | 1,295 (18.99%) | 438 (8.2%) | 84 (0.8%) |
| Unknown | 47 (1.7%) | 146 (2.8%) | 139 (7.62%) | 603 (8.84%) | 396 (7.5%) | 677 (6.5%) |
| **Chronic conditions** |  |  |  |  |  |  |
| Depression | 1,162 (42.2%) | 1,972 (37.5%) | 710 (38.95%) | 1,857 (27.22%) | 2,530 (47.6%) | 4,531 (43.6%) |
| Diabetes | 876 (31.8%) | 1,611 (30.6%) | 412 (22.60%) | 1,815 (26.61%) | 1,271 (23.9%) | 2,549 (24.5%) |
| Hypertension | 2,108 (76.5%) | 3,921 (74.5%) | 1,226 (67.25%) | 4,489 (65.81%) | 4,154 (78.2%) | 7,951 (76.6%) |
| Cancer | 852 (30.9%) | 1,414 (26.9%) | 455 (24.96%) | 1,219 (17.87%) | 1,668 (25.2%) | 3,028 (29.1%) |
| UFHealth IDR = University of Florida Health Integrated Data Repository<br>UTHealth Physicians CDW = University of Texas Health Science Center at Houston (UTHealth) Physicians Clinical Data Warehouse (CDW)<br>UMN AHC-IE CDR = University of Minnesota (UMN) Academic Health Center Information Exchange (AHC-IE) clinical data repository (CDR)<br>NHW = Non-Hispanic White<br>NHB = Non-Hispanic Black<br><sup>a</sup> The definitive AD cohort identified through the developed best performance CP rules<br><sup>b</sup> The potential AD cohort identified through the “having at least 1 AD related diagnosis code” criterion |  |  |  |  |  |  |

## DISCUSSION

In this study, we successfully developed and validated CP algorithms for identifying AD patients in EHRs, leveraging information from multiple EHR domains. Our study extended previous work of AD patient identification in several significant ways. First, in the development of the CP algorithm, we introduced more flexibility in the inclusion and exclusion criteria by considering and testing multiple EHR domains (i.e., medications, procedures, and keywords) in addition to AD/ADRD diagnosis codes. Second, in addition to considering information directly related with AD, we also considered diagnosis codes for cognitive decline. These codes may be recorded for patients with known AD status and including them in the CP algorithms further improved its coverage and robustness. Third, our final CP algorithm is simple (i.e., “*with at least 2 AD diagnosis, and with keywords in AD encounters*”), making it readily applied to other EHR systems. Further, compared with previous studies,^12–16^ our algorithm demonstrated superior performance, achieving higher sensitivity, while maintaining comparable PPV. Our final algorithm achieved a perfect sensitivity on the testing dataset, indicating that it can correctly identify all patients who truly have AD.

Nevertheless, there were a few false positives because: (1) in the unstructured data, patient is recorded as “*suspicious of two or more subtypes of ADRD*”, but was either diagnosed as “*having subtypes of ADRD other than AD*” or there is no conclusion yet at the time of our chart review; (2) in the unstructured data, patient was recorded as “*has dementia, possibly Alzheimer’s*”, but whether the patient truly has AD was not confirmed; and (3) potential document errors, where the patient had not been diagnosed with AD, but the condition was listed incorrectly in patient’s chart because of suspicious of AD. Our high-performing EHR-based CPs provides the opportunity of fast and accurate identification of AD patients from EHR systems, which can be used to build patient cohorts for research, clinical care, and public health initiatives. Our CP algorithms successfully identified a total of 2,756, 1,823, and 5,311 AD patients across the three sites, respectively, with most of them older than 65. Compared with the potential AD cohort, there was a higher representation of females in our final definitive AD cohort. This observation is consistent with previous reports suggesting that females are at higher risk of AD and other forms of dementia.^27–29^ Furthermore, we also observed higher percentages of NHBs and Hispanics compared with the potential AD patient population. Previous studies reported NHBs and Hispanics have higher rates of AD than NHWs.^30–32^ Moreover, the definitive AD cohort have higher prevalence of chronic diseases, including depression, diabetes, hypertension, and cancer.

Our study is not without limitations. One notable limitation is that when developing the CP algorithm, we only considered AD relevant keywords and did not consider the contexts in which the keywords are used within the unstructured clinical notes. For example, we did not account for negations (e.g., “*the patient does not have cognitive impairment*”), or references to people other than the patients themselves (e.g., “*he lived with a relative who has cognitive impairment*”). Advanced natural language processing methods may be used in future to further improve the accuracy of the CP algorithms.

In sum, we successfully developed and rigorously validated CP algorithms for accurately identifying AD patients. The final CP can be effectively applied in structured data alone, or in combination with unstructured clinical notes. The CPs we developed achieved good overall performance. The AD patient cohort identified through our CP can be used in downstream analysis to provide real-world evidence in understanding the disease burdens, social and behavioral determinants of health, patterns in utilization of services, and health outcomes in AD patients.

## Data Availability

All patient data used in the present study are sensitive and not publicly available.

## ACKNOWLEDGMENTS

None

## CONTRIBUTORSHIP STATEMENT

XH, ZC, QW, TL, SB and JB were responsible for the overall design, development, and evaluation of this study. XH, ZC, QW and JB did the initial drafts and revisions of the manuscript. JB was responsible for the conceptualization of the research. JT, LL, YZ, FW, HX, YC, RZ helped with the external validation of the algorithm. All authors reviewed the manuscript critically for scientific content, and all authors gave final approval of the manuscript for publication.

## CONSENT STATEMENT

This is a secondary analysis of electronic health records, where a waiver of consent was approved with an approved protocol by the University of Florida IRB.

